# Mimicking Clinical Trials with Synthetic Acute Myeloid Leukemia Patients Using Generative Artificial Intelligence

**DOI:** 10.1101/2023.11.08.23298247

**Authors:** Jan-Niklas Eckardt, Waldemar Hahn, Christoph Röllig, Sebastian Stasik, Uwe Platzbecker, Carsten Müller-Tidow, Hubert Serve, Claudia D. Baldus, Christoph Schliemann, Kerstin Schäfer-Eckart, Maher Hanoun, Martin Kaufmann, Andreas Burchert, Christian Thiede, Johannes Schetelig, Martin Sedlmayr, Martin Bornhäuser, Markus Wolfien, Jan Moritz Middeke

## Abstract

Clinical research relies on high-quality patient data, however, obtaining big data sets is costly and access to existing data is often hindered by privacy and regulatory concerns. Synthetic data generation holds the promise of effectively bypassing these boundaries allowing for simplified data accessibility and the prospect of synthetic control cohorts. We employed two different methodologies of generative artificial intelligence – CTAB-GAN+ and normalizing flows (NFlow) – to synthesize patient data derived from 1606 patients with acute myeloid leukemia, a heterogeneous hematological malignancy, that were treated within four multicenter clinical trials. Both generative models accurately captured distributions of demographic, laboratory, molecular and cytogenetic variables, as well as patient outcomes yielding high performance scores regarding fidelity and usability of both synthetic cohorts (n=1606 each). Survival analysis demonstrated close resemblance of survival curves between original and synthetic cohorts. Inter-variable relationships were preserved in univariable outcome analysis enabling explorative analysis in our synthetic data. Additionally, training sample privacy is safeguarded mitigating possible patient re-identification, which we quantified using Hamming distances. We provide not only a proof-of-concept for synthetic data generation in multimodal clinical data for rare diseases, but also full public access to synthetic data sets to foster further research.

**Graphical Abstract:** 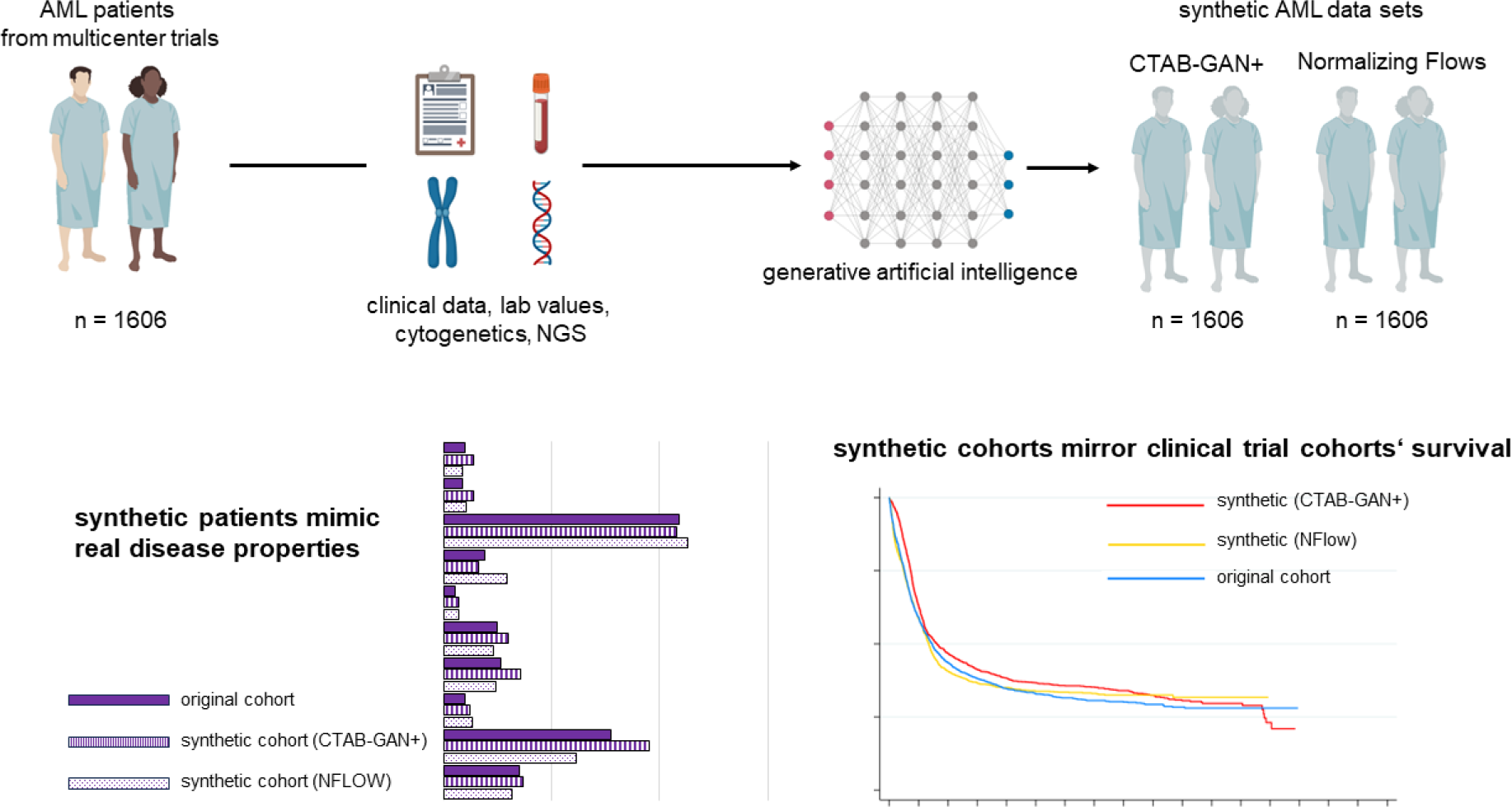

## Introduction

In the age of big data, the paucity of publicly available medical data sets is often staggering. Despite extensive data collection efforts, such as The Cancer Genome Atlas(1), the public availability of comprehensive entity-specific data sets remains largely unsatisfactory. Data sharing is often hindered by concerns of patient privacy, regulatory aspects, and proprietary interests.(2) These factors do not only impede progress in medical research but also establish a gatekeeping mechanism that restricts specific research inquiries to large institutions with access to extensive datasets. Collecting such data sets is a costly and time-consuming effort and especially later-phase clinical trials usually take years to complete and require millions in funding.(3,4) In particular, this is true for rare diseases, such as acute myeloid leukemia (AML), which is a genetically heterogenous and highly aggressive hematological malignancy with so far unsatisfactory patient outcomes despite recent advances in therapy.(5) In addition, the development of targeted therapies for defined subgroups leads to an increased need for control groups.(6) To gain insights into such burdensome malignant entities with unmet medical needs, a crowd-sourcing of data to refine risk stratification efforts and test treatment-related hypothesis is essential. If machine learning methods are to be deployed in such data sets, the size of available diverse training data is paramount for model robustness. Generative models, especially generative adversarial neural networks (GANs)(7), have exhibited remarkable capabilities in image generation(8), but can also effectively generate synthetic non-image data. The unique properties of generative artificial intelligence (AI) yield the prospect of synthesizing data based on real patients, which can be distributed at will since, ideally, synthetic data only mimics real patient data alleviating concerns of privacy. In this scenario, the synthetic data itself should preserve the biological characteristics of the disease under investigation to make inferences to real-world applications possible. At the same time, synthetic data should safeguard privacy of the underlying training cohort.

In this study, we employ two state-of-the-art technologies of generative modeling on a large training data set of four pooled multicenter clinical trials including AML patients with comprehensive clinical and genetic information. We investigate how closely the synthetic data resembles the real trial data aligning baseline characteristics and patient outcome. Further, we measure privacy conservation in the synthetic data. Additionally, we provide both final fully synthetic data sets comprising 1606 AML patients each in a publicly accessible repository to foster further research into this devastating disease.

## Methods

### Patient data

Multimodal clinical, laboratory, and genetic data (Table S1) were obtained from 1606 patients with non-M3 AML that were treated within previously conducted multicentric prospective clinical trials of the German Study Alliance Leukemia (SAL; AML96 [NCT00180115](9), AML2003 [NCT00180102](10), AML60+ [NCT00180167](11), and SORAML [NCT00893373](12)). Table S2 shows an overview of trial protocols. Eligibility was determined upon diagnosis of AML, age ≥18 years, and curative treatment intent. All patients gave their written informed consent according to the revised Declaration of Helsinki.(13) All studies were previously approved by the Institutional Review Board of the Technical University Dresden. Complete remission (CR), event-free survival (EFS), and overall survival (OS) were defined according to the revised ELN criteria.(14) Biomaterial was obtained from bone marrow aspirates or peripheral blood prior to treatment initiation. Next-Generation Sequencing (NGS) was performed using the TruSight Myeloid Sequencing Panel (Illumina, San Diego, CA, USA). Pooled samples were sequenced paired-end and a 5% variant allele frequency (VAF) mutation calling cut-off was used with human genome build HG19 as a reference as previously described in detail.(15) Additionally, high resolution fragment analysis for *FLT3*-ITD(16), *NPM1*(17), and *CEBPA*(18) was performed as described previously. For cytogenetics, standard techniques for chromosome banding and fluorescence-in-situ-hybridization (FISH) were used.

### Generative models

In our study, we used two state-of-the-art generative models exhibiting two fundamentally different concepts of data generation:

i. CTAB-GAN+(19) builds upon the Generative Adversarial Network (GAN)(20) architecture, consisting of two interlinked neural networks - the generator and the discriminator. These are jointly trained in an adversarial manner. The generator’s goal is to produce synthetic data that appears realistic, starting from random noise. In parallel, the discriminator seeks to differentiate between real and synthetic samples created by the generator. The training continues until the discriminator is no longer able to reliably distinguish real data from synthetic, indicating that the generator has successfully approximated the distribution of the real data.
ii. Normalizing Flows (NFlow)(21) presents an alternative approach for synthesizing data from complex distributions. This comprises a sequence of invertible transformations, starting from a simple base distribution. Each transformation, or ’flow’, gradually modifies this base distribution into a more complex one that better mirrors the actual data. Importantly, these transformations are stackable, meaning they can be applied successively to incrementally increase the complexity of the modeled distribution. All parameters defining these flows are learned directly from the data, allowing the model to accurately capture the underlying data distribution. Note, that we used a modification of NFlow for survival data provided by the Synthcity(22) software framework.

No imputation of missing data was performed in the original data set, thus both final synthetic data sets also contain missing data to adequately represent real-world conditions. Hyperparameter tuning was performed using the Optuna framework allowing both generative models to capture the best possible representation of the original data. Afterwards, we trained each model with five different random seeds and sampled from it three times, which generated 15 synthetic datasets for each model. Results are reported for each highest-performing synthetic data set, respectively.

### Evaluation of synthetic data performance

To assess the fidelity und usability of synthetic data, previously proposed evaluation metrics were used to provide a comprehensive overview of model performance. In particular, Basic Statistical Measure, Regularized Support Coverage, and Log-transformed Correlation Score were used to evaluate the fidelity of the data in general via our implementation based on the descriptions by Chundawat et al.(23). The second set of metrics – Kaplan-Meier-Divergence, Optimism and Short-Sightedness - was previously introduced by Norcliffe et al.(24) for synthetic survival data, and implemented in Synthcity(22). For improved comparability, performance metrics were normalized on a scale from 0 (inadequate representation of original data) to 1 (optimal representation). An overview of the underlying methodologies of these metrics is provided in Table S3. For detailed information, we refer the interested reader to the original publications.(23,24)

### Assessment of privacy conservation

To assess potential privacy implications of synthetic data, we customized the method proposed by Platzer and Reutterer(25) to accommodate for smaller sample sizes. We partitioned the original training data (80% of total) into four subsets, matching the size of the test dataset (20%) for balanced comparisons (Fig. S1). Calculations were performed using Hamming distance(26) for categorical features. Numerical variables were binned (n=10 bins each) and thereby categorized to enable Hamming distance calculations. Given the nature of the Hamming distance metric, the average minimum distance effectively denotes the number of variables that would need to be altered for a synthetic patient to match a real patient. We compared the average distances of the synthetic data to the training (syn **→** train) and test sets (syn **→** test). The relationship between both can be expressed as a coefficient for each synthetic data set compared to training and test set:

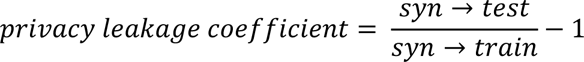

By analyzing whether the synthetic data is closer to the training set compared to the test set, we can assess whether the synthetic data is overly representative of the training data, thereby posing potential privacy concerns. If the average distances from the synthetic data to the training and test data are equally small, the privacy leakage coefficient will also be small. The lower the privacy leakage coefficient, the lower the likelihood of re-identification for patients in the training set. We assumed that values above 0.05 signal potential privacy breaches, as they suggest the synthetic data is substantially closer to the training set than to the test set. Conversely, values below 0.05 denote a favorable privacy safeguard, signaling similar distances between the training and test sets. Additionally, the number of exact subject matches between the synthetic and original cohorts was determined.

### Statistical analysis

Pairwise analyses were conducted between the original and both synthetic data sets. Normality was assessed using the Shapiro-Wilk test. If the assumption of normality was met, continuous variables between two samples were analyzed using the two-sided unpaired t-test. If the assumption of normality was violated, continuous variables between two samples were analyzed using the Wilcoxon rank sum (syn. Mann-Whitney) test. Fisher’s exact test was used to compare categorical variables. Univariate analyses for binary outcomes (CR rate) were carried out via logistic regression to obtain odds ratios (OR) and 95% confidence intervals (95%-CI). Time-to-event analyses (EFS, OS) were carried out using Cox proportional hazard models to obtain hazard ratios (HR) and 95%-CI. Kaplan-Meier analyses were performed for time-to-event data (EFS, OS) and corresponding log-rank tests are reported. Median follow-up time was calculated using the reverse Kaplan-Meier method.(27) All tests were carried out as two-sided tests. Statistical significance was determined using a significance level α of 0.05. Statistical analysis was performed using STATA BE 18.0 (Stata Corp, College Station, TX, USA).

### Data availability

The final synthetic data sets generated and analyzed for the purpose of this study are publicly available at https://zenodo.org/record/8334265

### Code availability

The code generated for the purpose of this study is publicly available at https://github.com/waldemar93/synthetic_data_pipeline

## Results

### Synthetic cohorts generated by CTAB-GAN+ and NFlow score highly in fidelity metrics

We generated equally sized data sets of n=1606 synthetic patients with each generative model to compare patient variables to the original cohort. The fidelity of synthetic data was assessed with three previously proposed performance metrics scaled from 0 (inadequate representation) to 1 (optimal representation). First, the distribution of each individual variable was compared between original and synthetic data again yielding high scores for both models (Regularized Support Coverage(23) for CTAB-GAN+: 0.95 and NFlow: 0.97). Second, continuous numerical variables were assessed by comparing mean, median, and standard deviation between original and synthetic data per variable (Basic Statistical Measure(23)) showing high scores for both CTAB-GAN+ (0.91) and NFlow (0.92). Third, regarding accurate representations of inter-variable correlations, CTAB-GAN+ and NFlow achieved a Log-Transformed Correlation Score(23) of 0.75 and 0.74, respectively. An overview of performance metrics is provided in Tab. S4 (usability; survival metrics are reported with survival analysis).

### Synthetic clinical and genetic patient characteristics closely mimic those of real patients

Baseline patient characteristics compared between real and synthetic patients are shown in Table 1. It has to be noted that given the large sample sizes (three groups with n=1606 each), even small effect sizes yield statistically significant differences. For instance, median age in the original cohort was 56 years, while synthetic patients generated by CTAB-GAN+ had a slightly younger median age of 53 years (*p*=0.0001), whereas NFlow-generated patients had a slightly older median age of 58 years (*p*=0.039). Sex distribution did not differ between NFlow and the original cohort, while CTAB-GAN+ generated more males than females (NFLOW: 56.2% vs. 43.8%; original: 52.2% vs. 47.8%; *p*=0.023). The rates of *de novo*, secondary, and therapy-associated AML did not differ significantly for CTAB-GAN+ generated patients, while NFlow generated fewer *de novo* and more therapy-associated AML patients compared to the original cohort. Hemoglobin levels and platelet count did not differ significantly between the original and the synthetic cohorts, while synthetic patients generated by CTAB-GAN+ showed a significantly higher median white blood cell count than the original cohort.

**Table 1.** Distribution of baseline characteristics between the original and synthetic cohort. Boldface indicates statistical significance (*p* < 0.05). *p*-values are calculated using two-sample comparisons between each of the synthetic cohorts and the baseline cohort for reference. Abbreviations: Hb: hemoglobin; IQR: interquartile range; n: number; Plt: platelet count; WBC: white blood cell count.

| clinical data | original cohort | CTAB-GAN+ | *p* | NFlow | *p* |
| --- | --- | --- | --- | --- | --- |
| number of patients | 1606 | 1606 |  | 1606 |  |
| age, median (IQR) | 56 (44 - 65) | 53 (42 - 64) | **0.0001** | 58 (47 – 66) | **0.039** |
| sex, n (%) |  |  | **0.023** |  | 0.672 |
| female | 768 (47.8) | 703 (43.8) |  | 781 (48.6) |  |
| male | 838 (52.2) | 903 (56.2) |  | 825 (51.4) |  |
| AML status, n (%) |  |  |  |  |  |
| de novo | 1339 (83.4) | 1339 (83.4) | 1.000 | 1250 (77.8) | **0.041** |
| secondary | 195 (12.1) | 193 (12.0) | 0.914 | 200 (12.5) | 0.554 |
| therapy-associated | 54 (3.4) | 57 (3.5) | 0.847 | 83 (5.2) | **0.007** |
| extramedullary disease, n (%) | 224 (13.9) | 228 (14.2) | 0.409 | 279 (17.4) | **0.003** |
| **laboratory values** |  |  |  |  |  |
| WBC, median (IQR) in GPt/l | 19.5 (4.5 - 53.4) | 27.0 (8.3 - 69.6) | **<0.0001** | 14.4 (5.8 – 55.3) | 0.832 |
| Hb, median (IQR) in mmol/l | 5.9 (5.0 - 8.6) | 5.8 (5.0 - 7.0) | 0.949 | 5.9 (5.2 – 6.8) | 0.988 |
| Plt, median (IQR) in GPt/l | 50.0 (27.0 – 94.0) | 49.7 (31.0 - 93.4) | 0.073 | 48.0 (26.2 – 94.5) | 0.405 |

Fifty molecular and cytogenetic alterations were included in generating synthetic patients. Figure 1 displays the distribution of these alterations across the original and synthetic cohorts (absolute numbers and *p*-values are provided in Tab. S5). These alterations encompass genes that code for epigenetic regulators (Fig. 1A), the cohesin complex (Fig. 1B), transcription factors (Fig. 1C), *TP53* and *Nucleophosmin 1* (Fig. 1D), signaling factors (Fig. 1E), components of the spliceosome (Fig. 1F), and cytogenetic aberrations with established impact on patient outcome (Fig. 1G). Overall, the rates of alterations in both synthetic cohorts were in a plausible range with a few deviations from the original cohort of high statistical significance, such as NFlow-generated frequencies of *BCORL1, DNMT3A, PHF6*, and *ZRSR2,* as well as CTAB-GAN+-generated frequencies of *CUX1* and *GATA2* while the remainder of alterations showed only negligible differences. Aside from the frequency per individual alteration, the co-occurrences of alterations play an important role in disease biology, which should be also captured in high-quality synthetic data. Fig. 2 shows the relative differences between the original cohort and CTAB-GAN+ (Fig. 2A) and NFlow (Fig. 2B) regarding co-occurring mutations. We found high congruencies for co-occurrences compared to the original cohort, while deviations were commonly found in alterations that had a low frequency in the original cohort.

**Figure 1.**
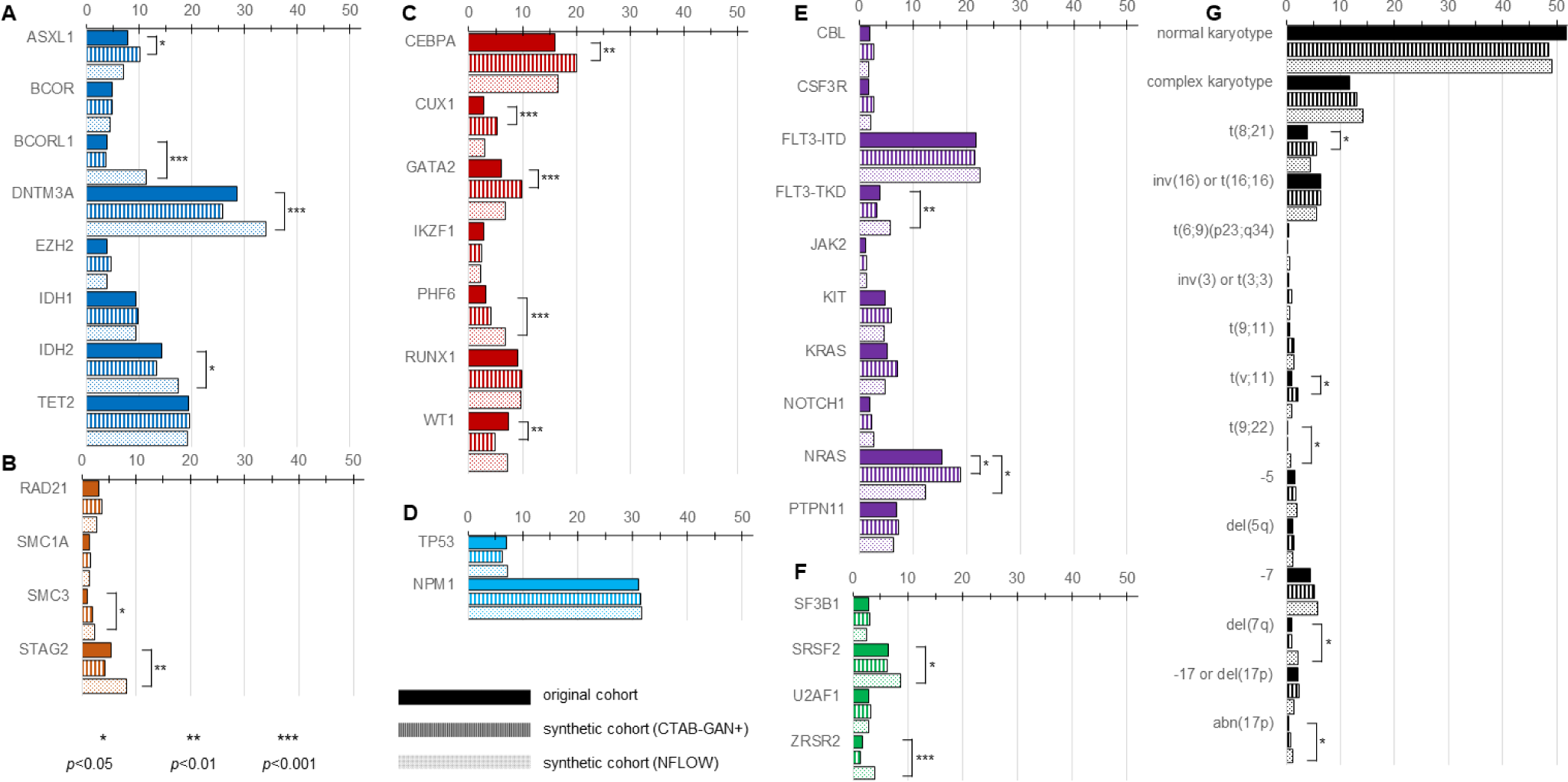
Distribution of molecular and cytogenetic alterations between real and synthetic patients. 50 molecular genetic and cytogenetic alterations were included in generative modeling. Molecular genetics were originally assessed by next-generation sequencing using a targeted myeloid panel including genes that encode for epigenetic regulators (A, dark blue), the cohesion complex (B, orange), transcription factors (C, red), NPM1 and TP53 (D, light blue), signaling factors (E, purple), and the spliceosome (F, green). Cytogenetic aberrations (G, black) were selected based on previously demonstrated impact on patient outcomes. Distributions for all variables are denoted as percentages of each respective cohort. Overall, both synthetic cohorts well represented the distribution of alterations in the original cohort with only slight deviations denoted by highly statistically significant (*p*<0.001) differences in *BCORL1, DNMT3A, PHF6*, and *ZRSR2* for NFlow, as well as *CUX1* and *GATA2* for CTAB-GAN+.

**Figure 2.**
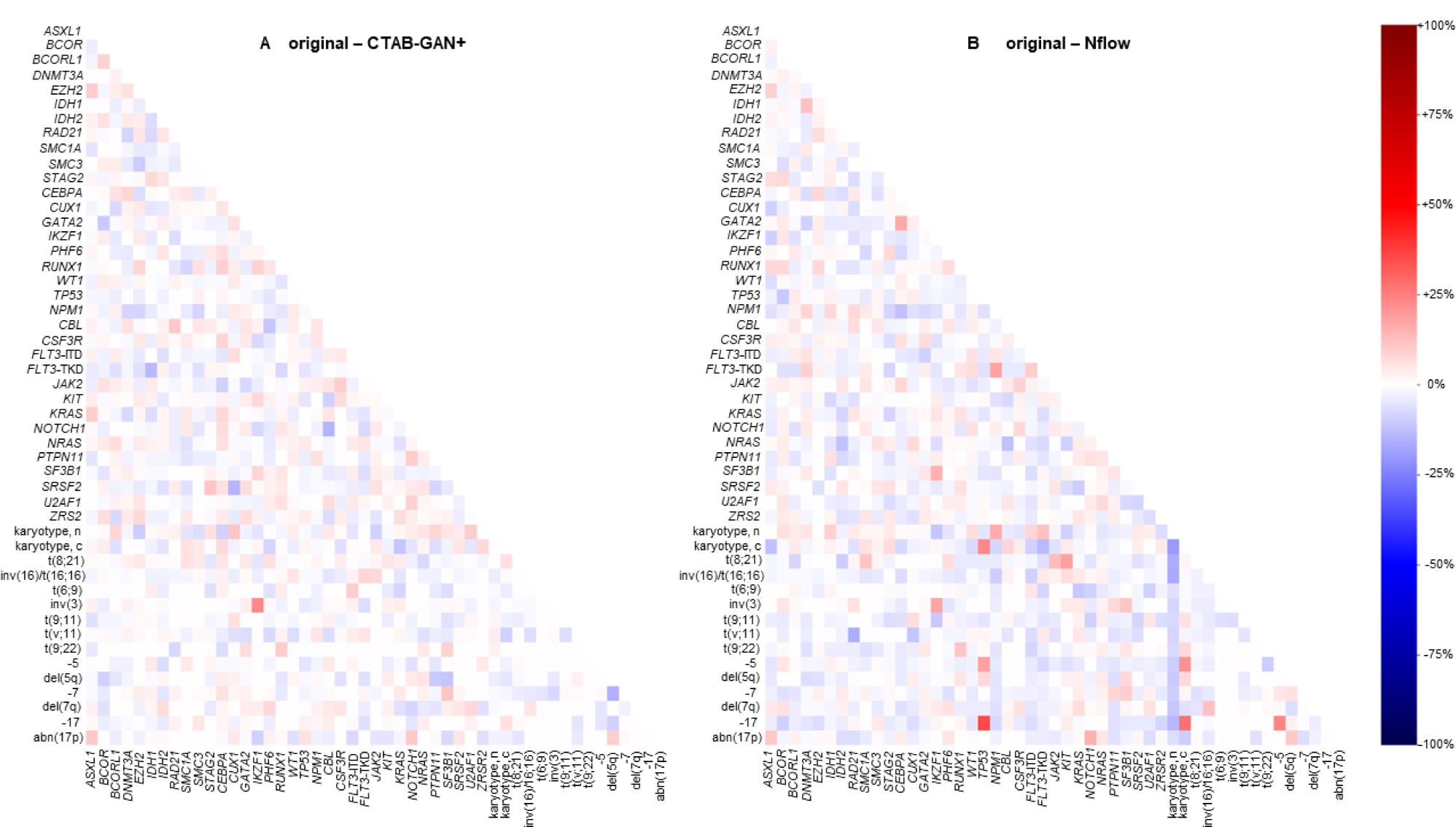
Heatmaps for relative differences of genetic associations. The difference in co-occurrences of genetic alterations are plotted. Relative increases (red) or decreases (blue) are displayed on a scale from −100% to + 100%. The overlap between the original cohort and CTAB-GAN+ (A), as well as original and NFlow (B) showed high congruency. Increases or decreases in co-occurring genetic alterations were commonly found to affect alterations with low frequency in the original cohort.

### Synthetic cohorts match real patients in outcome and survival analysis

Median follow-up for the original cohort was 89.5 months (95%-CI: 85.5-95.4). The synthetic cohorts had a median follow-up of 91.3 months (CTAB-GAN+, 95%-CI: 84.8-98.0) and 74.3 months (NFlow, 95%-CI: 70.9-77.4). Table 2 shows a detailed comparison of patient outcome between the original and both synthetic cohorts. For CR rates, we found no significant differences between the original (70.7%) and both synthetic cohorts (CTAB-GAN+: 73.7%; NFlow: 69.1%). Median EFS in the original cohort was 7.2 months while both CTAB-GAN+ with 12.8 months and NFlow with 9.0 months deviated with high significance. This effect can arguably be attributed to both CR rate and OS being included in hyperparameter tuning, while EFS was exempt from hyperparameter tuning. Kaplan-Meier analysis nevertheless showed a plausible representation of the survival curves for both synthetic cohorts regarding EFS (Fig. 3A). Median OS for the original cohort was 17.5 months while the CTAB-GAN+cohort had a median OS of 19.5 months (p<0.0001) and NFlow of 16.2 months (p=0.055). Kaplan-Meier analysis (Fig. 3B) showed similar behavior of survival curves as for EFS. This was also evident with regard to usability metrics for synthetic survival data introduced by Norcliffe et al.(24): We found both CTAB-GAN+ and NFlow to score high in our test set with normalized performance results (+1 is optimal representation, 0 is inadequate representation, Tab. S4). Kaplan-Meier-Divergence, i.e. the degree to which survival curves of synthetic and real data differ, was low for both synthetic data sets (CTAB-GAN+: 0.97, NFlow: 0.98). Neither model showed overt optimism or overt pessimism in representing survival data (CTAB-GAN+: 0.98, NFlow: 0.99). For both EFS and OS, the curve of CTAB-GAN+ showed no stabilization of survival rates towards the end of the follow-up period in comparison to the curve of the original cohort while NFlow tends to censor a higher rate of patients after passing the 2-year follow-up mark. Nonetheless, Short-sightedness, i.e. failure to predict beyond a certain time point, was also low for both models, however slightly favoring CTAB-GAN+ over NFlow (CTAB-GAN+: 0.99, NFlow: 0.93) arguably corresponding to the censoring tendency of NFlow.

**Figure 3.**
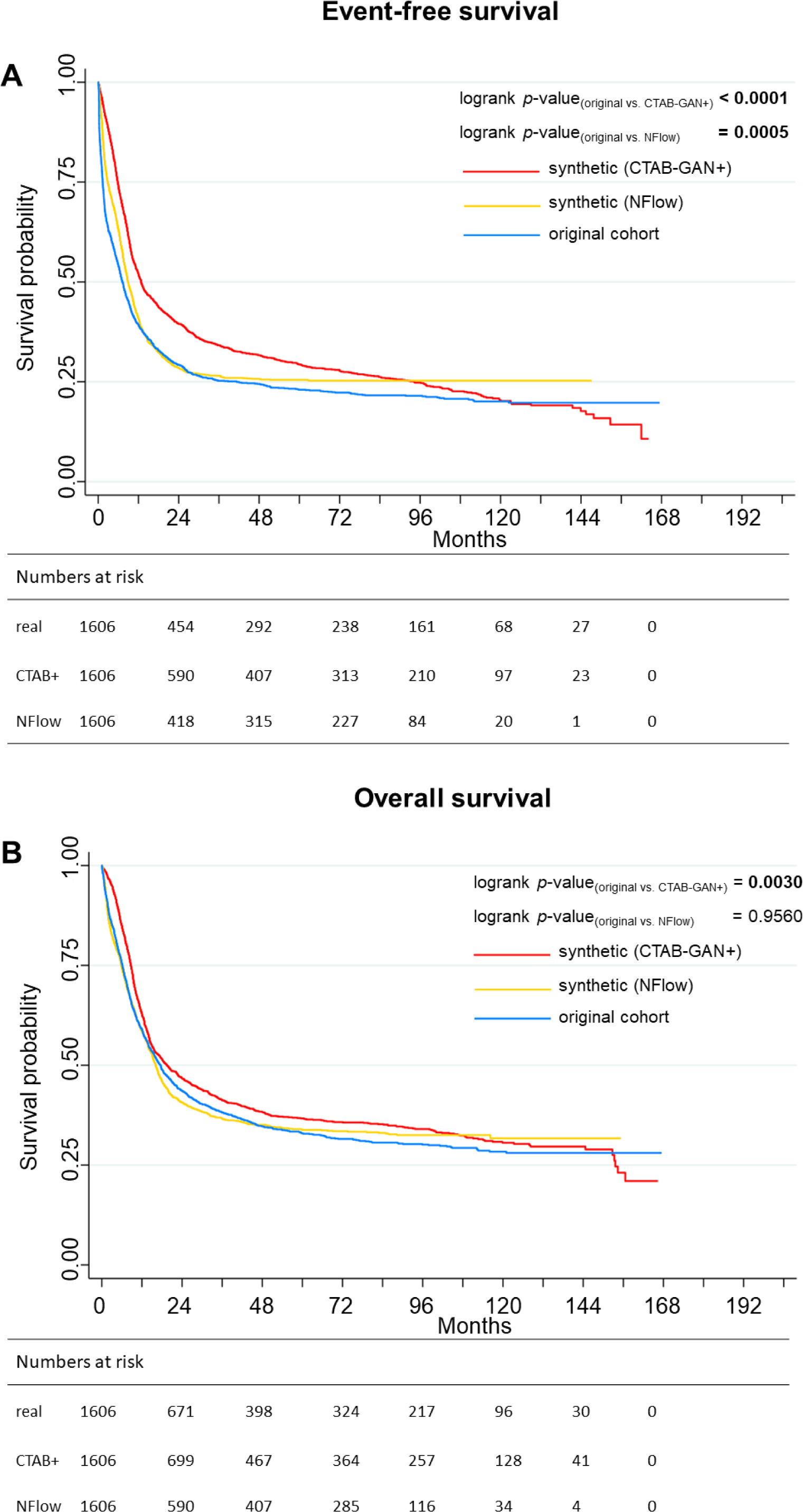
Comparison of survival curves between original and synthetic cohorts. Event-free survival (EFS) deviated significantly from the original cohort for both synthetic cohorts (A). For the NFlow-generated cohort, there was no significant deviation from the original distribution for overall survival (OS), while the CTAB-GAN+-generated cohort again differed significantly (B). Interestingly, while the survival curve for CTAB-GAN+ displays a plausible curve up until ten years of follow-up, the curve shows no stabilization of survival rates in the end as the original cohort does. Contrastingly, the survival curve for NFlow shows an overall plausible course, however, NFlow tends to overtly censor patients after two years of follow-up.

**Table 2.**
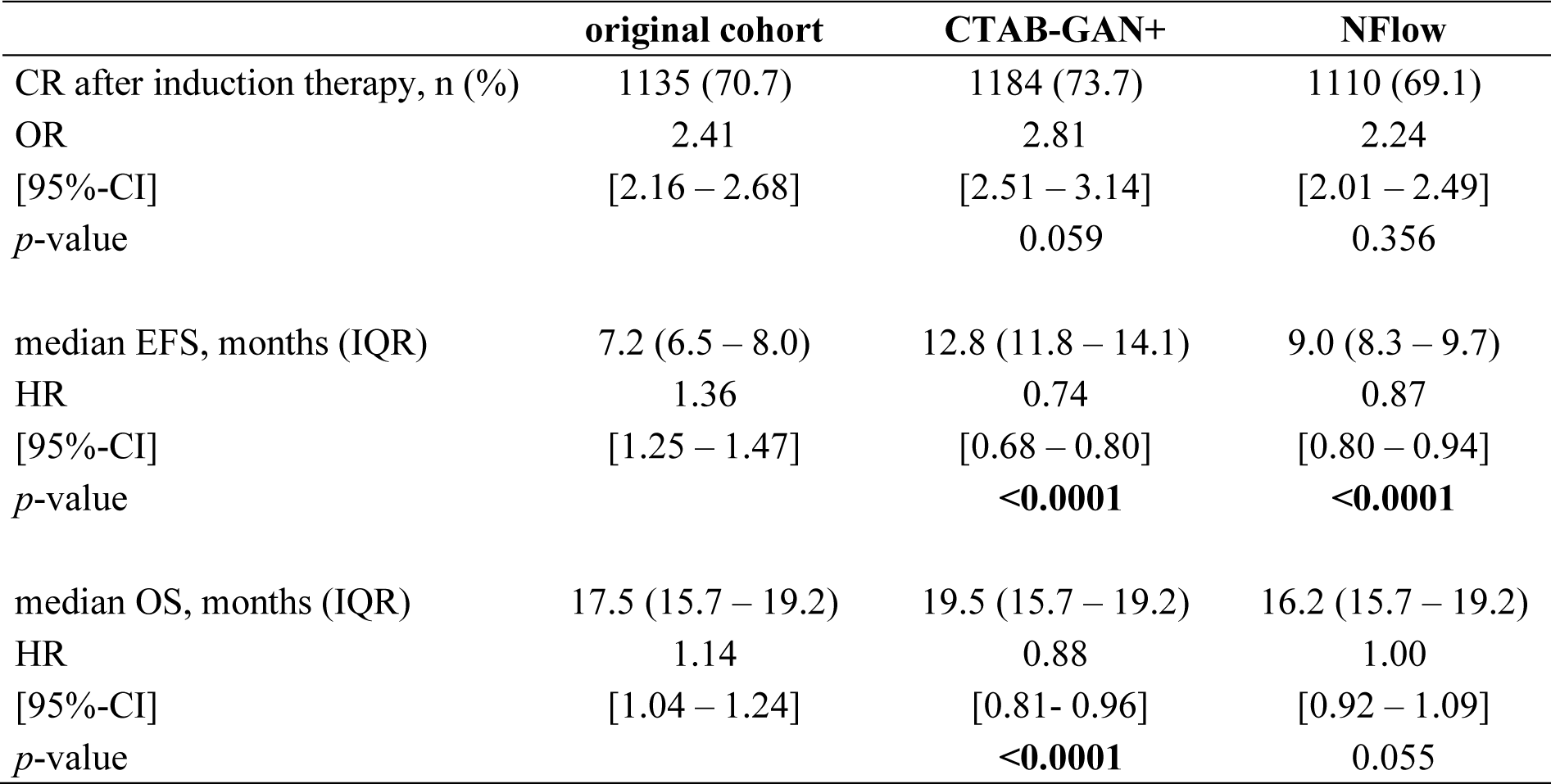
Comparison of patient outcomes between the original and synthetic cohort. Logistic regression and Cox proportional hazard models were used to obtain odds ratios (OR) for achievement of complete remission (CR) and hazard ratio (HR) with corresponding 95%-confidence intervals (95%-CI). Boldface indicates statistical significance (*p* < 0.05). *p*-values are calculated using two-sample comparisons between each of the synthetic cohorts and the original cohort for reference. Other abbreviations: n: number.

|  | original cohort | CTAB-GAN+ | NFlow |
| --- | --- | --- | --- |
| CR after induction therapy, n (%) | 1135 (70.7) | 1184 (73.7) | 1110 (69.1) |
| OR | 2.41 | 2.81 | 2.24 |
| [95%-CI] | [2.16 – 2.68] | [2.51 – 3.14] | [2.01 – 2.49] |
| $p$-value | | 0.059 | 0.356 |
| median EFS, months (IQR) | 7.2 (6.5 – 8.0) | 12.8 (11.8 – 14.1) | 9.0 (8.3 – 9.7) |
| HR | 1.36 | 0.74 | 0.87 |
| [95%-CI] | [1.25 – 1.47] | [0.68 – 0.80] | [0.80 – 0.94] |
| $p$-value | | **<0.0001** | **<0.0001** |
| median OS, months (IQR) | 17.5 (15.7 – 19.2) | 19.5 (15.7 – 19.2) | 16.2 (15.7 – 19.2) |
| HR | 1.14 | 0.88 | 1.00 |
| [95%-CI] | [1.04 – 1.24] | [0.81- 0.96] | [0.92 – 1.09] |
| $p$-value | | **<0.0001** | 0.055 |

### Synthetic data captures risk associations of individual variables for explorative analyses

In order to be useful for explorative analyses, synthetic data needs to recapitulate risk associations of individual variables. The ELN2022 recommendations represent one of the most widely used guidelines for risk stratification.(14) Hence, previously established markers of favorable (normal karyotype, t(8;21), inv(16) or t(16;16) mutations of *NPM1*, *CEBPA*-bZIP in frame mutations), intermediate risk (*FLT3*-ITD, t(9;11)), or adverse risk (complex karyotype, −5, del(5q), −7, −17, mutations of *TP53, RUNX1, ASXL1*), and age were evaluated using univariable analyses per cohort for their impact on achievement of CR, EFS, and OS. All effects for achievement of CR, EFS, and OS showed the same directionality – favorable affects in the original cohort were also favorable in synthetic cohorts and *vice versa* – and significance – effects that were significant in the original cohort were also significant in synthetic cohorts and *vice versa* (except for del(5q) being significantly associated with failure to achieve CR in the original cohort while this effect turned out to be non-significant in the NFlow-generated cohort). Importantly, no inverse effects – a variable that would be favorable in the original cohort would be adverse in a synthetic cohort or *vice versa* – were observed. Detailed outcomes per variable are reported for CR (Tab. S6), EFS (Tab. S7), and OS (Tab. S8).

### Synthetically generated cohorts safeguard real patient data and prohibit re-identification

Privacy conservation was measured by: i) number of exact matches between original and synthetic cohorts, ii) a privacy leakage coefficient based on Hamming distance, and iii) absolute Hamming distances showing the number of variables to be altered per synthetic patient to match a real patient. First, for both synthetic data sets the number of exact matches compared to the original cohort was zero. Second, the average minimum distances compared between datapoints in training and test sets were similar for the original cohort, as well as synthetic data from both CTAB-GAN+ and NFlow (Tab. 3). The privacy leakage coefficient – the quotient of Hamming distances between synthetic to test divided by synthetic to training data where small values (<0.05) indicate a small difference between the distances of synthetic data to training and test data, and therefore, indicate no privacy breach – was very low for both CTAB-GAN+ and NFlow (Tab. 3). This signals a low likelihood of re-identification for both synthetic datasets. Third, the median number of variables that would have to be altered to assign a synthetic patient to a training set patient was nine for both CTAB-GAN+ and NFlow.

**Table 3.**
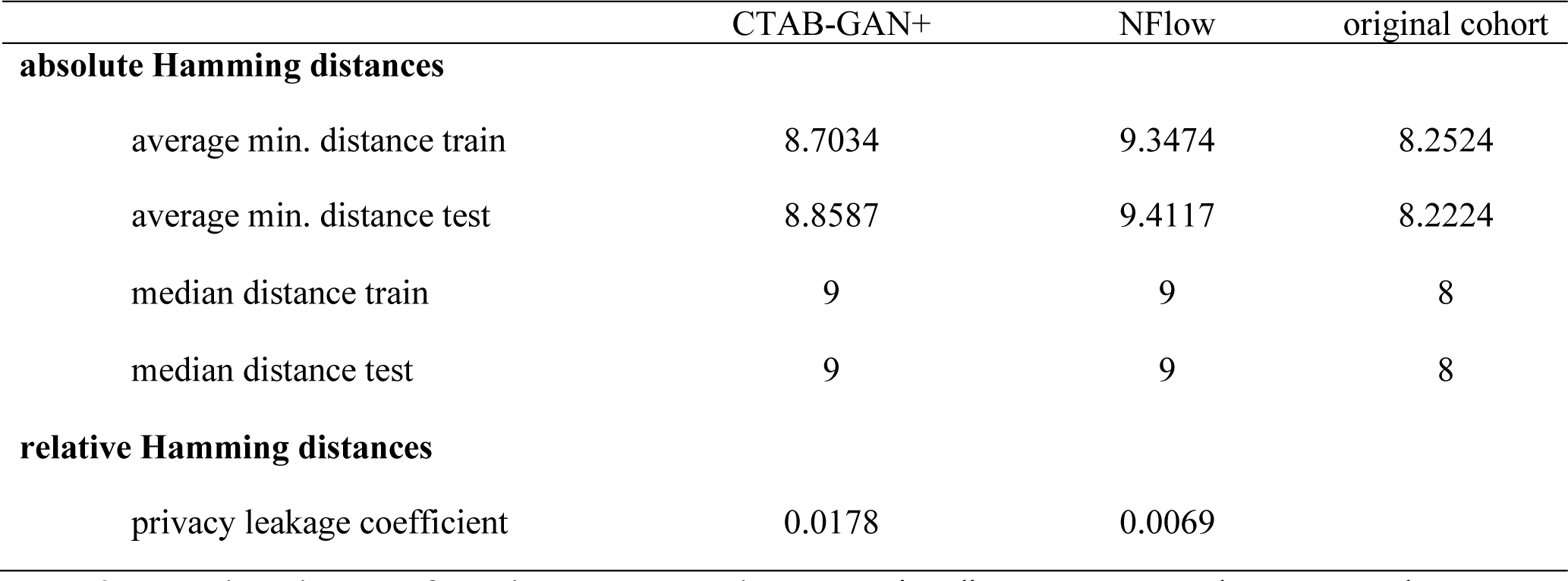
Hamming distances for privacy conservation. Hamming distances were used to measure the distance between two points within and between equally sized subsets of training (four sets of 20%) and test data (20%). The median distance represents the number of variables that have to be altered (and matched exactly) to fit a real patient. A threshold for the privacy leakage coefficient of 0.05 for relative distances was set where values above 0.05 signal potential privacy breaches. Both synthetic data sets fell well below the 0.05 threshold signaling larger distances between synthetic and training data, which make a re-identification of training set patients unlikely.

## Discussion

Synthetic data provide an attractive solution to circumvent issues in current standards of data collection and sharing. These issues encompass first and foremost the time- and cost-intensive data collection process that usually involves enrollment of patients in prospective clinical trials presenting ever-increasing costs both regarding funding and time until completion, as well as ethical concerns inherent in clinical research with human subjects.(3,4) The prospect of using synthetic data as a novel kind of control group in prospective trials while effectively alleviating the need to enroll a larger number of patients and cutting costs bears the question of how closely such synthetic control arms match real-world cohorts. We used two generative AI technologies, a state-of-the-art GAN, CTAB-GAN+, and NFlow, to mimic the distribution of patient variables from four different previously conducted prospective multicenter trials including a total of 1606 patients with AML. Both models demonstrated high performance in previously established evaluation metrics that assess fidelity and usability of synthetic tabular data.(23,24) The comparison of distributions per variable between original and real data further showed close resemblances. Notably, even for statistically significant deviations from the original cohort, differences in effect sizes (e.g. age difference, difference in rates of occurrence for genetic alterations etc.) were often small. Inherent to hypothesis testing with such large sample sizes, even clinically irrelevant deviations can yield statistically significant differences. Importantly, inter-variable relationships were conserved in synthetic data: In univariable analyses both effect direction and statistical significance was well captured by both generative models effectively enabling explorative investigations in such data sets.

Once real data is obtained, privacy concerns often inhibit public access and thus impede data sharing and third-party hypothesis testing. Frequently used practices range from de-identifying or anonymizing data to more advanced computational approaches. De-identification or anonymization (e.g. removing names and birth dates), as well as adding artificial noise to the original data have recently been proven to be unsafe in terms of guarding privacy as reidentification attacks can successfully unveil patients’ identity.(28–30) Computational advances in both federated(31) and swarm learning(32) where machine learning models are trained across multiple locations and only either models or weights are shared rather than the data itself provide a viable alternative. Nevertheless, these technologies are vulnerable to data reconstructions, e.g. via data leakage from model gradients.(33–35) Inherent to synthetic data generation in terms of privacy safeguards is a trade-off between usability and privacy where an increase in each negatively affects the other.(36) Ideally, synthetic data should not be re-identifiable but at the same time closely match the original distributions. Zero exact matches were observed in our synthetic cohorts. Additionally, Hamming distances showed that reconstruction of original training samples is highly unlikely given the number of variables per synthetic patient that would have to be altered in order to match a training cohort patient.

The generation of synthetic data is, as all machine learning models are, fundamentally limited by the data that the model is trained on. This implies that external users should be aware of the properties of the training data that went into the generation of a synthetic data set in order to either select the right data set for their research question or *vice versa*, adapt the research question to the available data. It is therefore important to note, that patients in our trials have all been treated with intensive anthracycline-based therapy and largely stem from a Middle-European ethnic background. Hence, our generated synthetic AML data sets may not fully capture features of other populations let alone other treatment modalities, such as less intensive therapy or targeted agents. The incorporation of these modalities will be addressed in future works. Since ML models thrive on large and diverse data sets, synthetic data generation from medical records is caught in a paradoxical loop: Available data is sparse, synthetic data can potentially accommodate for sparse available real data, synthetic data requires large and diverse sets of real data to meaningfully represent the population.(37) Therefore, the generation of synthetic data is likely more robust, if training data from large multicenter cohorts is used. Nonetheless, the availability of synthetic data promises a democratization of clinical research. In similar efforts, Azizi et al.(38) and D’Amico et al.(39) explored synthetic data generation in cancer. Azizi et al.(38) used data from a previously conducted clinical trial in colorectal cancer to generate synthetic data using conditional decision trees. Focusing on myelodysplastic neoplasms (MDS), D’Amico et al.(39) used a conditional Wasserstein tabular GAN to generate synthetic MDS patients from the GenoMed4All database. Both groups conclude the feasibility of either method to generate synthetic data that closely resemble the original data distributions and provide access to their synthetic data. Such studies may alleviate a common gatekeeping mechanism of costly data collection efforts that are often restricted to large well-funded medical centers. Further, this also extends to cross-domain applications involving medical data, e.g. the training of a ML model by a third party that requires large sets of training data.

In summary, we demonstrate the feasibility of two different technologies of generative AI to create synthetic clinical trial data that both closely mimic disease biology and clinical behavior, as well as conserve the privacy of patients in the training cohort. Generating such large synthetic data sets based on multicenter clinical trial training data holds the promise of enabling a new kind of clinical research improving upon data accessibility, while ameliorating current hindrances in data sharing.

## Acknowledgements

We thank all contributing physicians, laboratories, and nurses associated with the German Study Alliance Leukemia and especially participating patients for their valuable contributions.

## Authorship Contributions

J.-N.E., W.H., M.W., and J.M.M. designed the study. J.-N.E., C.R, U.P., C. M.-T., H.S., C.D.B., C.S., K.S-E., M.H., M.K., A.B., C.T., J.S., M.B., and J.M.M. provided patient samples. S.S. and C.T. performed molecular analysis. W.H. trained generative models. J.-N.E. performed statistical analysis and wrote the initial draft. All authors had access to all of the data, analyzed the data, provided critical scientific insights and revised the draft. All authors agreed to the final version of the manuscript and the decision to submit it for publication.

## References

1. The Cancer Genome Atlas Program - National Cancer Institute [Internet]. 2018 [cited 2020 Sep 1]. Available from: https://www.cancer.gov/about-nci/organization/ccg/research/structural-genomics/tcga

2. Taitsman JK, Grimm CM, Agrawal S. Protecting Patient Privacy and Data Security. New England Journal of Medicine. 2013 Mar 14;368(11):977–9.

3. Stewart DJ, Stewart AA, Wheatley-Price P, Batist G, Kantarjian HM, Schiller J, et al. The importance of greater speed in drug development for advanced malignancies. Cancer Med. 2018 Mar 30;7(5):1824–36.

4. Martin L, Hutchens M, Hawkins C, Radnov A. How much do clinical trials cost? Nature Reviews Drug Discovery. 2017 Jun 1;16(6):381–2.

5. Döhner H, Weisdorf DJ, Bloomfield CD. Acute Myeloid Leukemia. New England Journal of Medicine. 2015 Sep 17;373(12):1136–52.

6. Estey E, Othus M, Gale RP. New study-designs to address the clinical complexity of acute myeloid leukemia. Leukemia. 2019 Mar;33(3):567–9.

7. Goodfellow IJ, Pouget-Abadie J, Mirza M, Xu B, Warde-Farley D, Ozair S, et al. Generative Adversarial Networks [Internet]. arXiv; 2014 [cited 2022 Jul 21]. Available from: http://arxiv.org/abs/1406.2661

8. Kazeminia S, Baur C, Kuijper A, van Ginneken B, Navab N, Albarqouni S, et al. GANs for medical image analysis. Artificial Intelligence in Medicine. 2020 Sep 1;109:101938.

9. Röllig C, Thiede C, Gramatzki M, Aulitzky W, Bodenstein H, Bornhäuser M, et al. A novel prognostic model in elderly patients with acute myeloid leukemia: results of 909 patients entered into the prospective AML96 trial. Blood. 2010 Aug 12;116(6):971–8.

10. Schaich M, Parmentier S, Kramer M, Illmer T, Stölzel F, Röllig C, et al. High-dose cytarabine consolidation with or without additional amsacrine and mitoxantrone in acute myeloid leukemia: results of the prospective randomized AML2003 trial. J Clin Oncol. 2013 Jun 10;31(17):2094–102.

11. Röllig C, Kramer M, Gabrecht M, Hänel M, Herbst R, Kaiser U, et al. Intermediate-dose cytarabine plus mitoxantrone versus standard-dose cytarabine plus daunorubicin for acute myeloid leukemia in elderly patients. Ann Oncol. 2018 01;29(4):973–8.

12. Röllig C, Serve H, Hüttmann A, Noppeney R, Müller-Tidow C, Krug U, et al. Addition of sorafenib versus placebo to standard therapy in patients aged 60 years or younger with newly diagnosed acute myeloid leukaemia (SORAML): a multicentre, phase 2, randomised controlled trial. Lancet Oncol. 2015 Dec;16(16):1691–9.

13. World Medical Association. World Medical Association Declaration of Helsinki: Ethical Principles for Medical Research Involving Human Subjects. JAMA. 2013 Nov 27;310(20):2191–4.

14. Döhner H, Wei AH, Appelbaum FR, Craddock C, DiNardo CD, Dombret H, et al. Diagnosis and Management of AML in Adults: 2022 ELN Recommendations from an International Expert Panel. Blood. 2022 Jul 7;blood.2022016867.

15. Stasik S, Schuster C, Ortlepp C, Platzbecker U, Bornhäuser M, Schetelig J, et al. An optimized targeted Next-Generation Sequencing approach for sensitive detection of single nucleotide variants. Biomol Detect Quantif. 2018 May;15:6–12.

16. Thiede C, Steudel C, Mohr B, Schaich M, Schäkel U, Platzbecker U, et al. Analysis of FLT3-activating mutations in 979 patients with acute myelogenous leukemia: association with FAB subtypes and identification of subgroups with poor prognosis. Blood. 2002 Jun 15;99(12):4326– 35.

17. Thiede C, Koch S, Creutzig E, Steudel C, Illmer T, Schaich M, et al. Prevalence and prognostic impact of NPM1 mutations in 1485 adult patients with acute myeloid leukemia (AML). Blood. 2006 May 15;107(10):4011–20.

18. Taube F, Georgi JA, Kramer M, Stasik S, Middeke JM, Röllig C, et al. CEBPA Mutations in 4708 Patients with Acute Myeloid Leukemia - Differential Impact of bZIP and TAD Mutations on Outcome. Blood. 2021 Jul 28;blood.2020009680.

19. Zhao Z, Kunar A, Birke R, Chen LY. CTAB-GAN+: Enhancing Tabular Data Synthesis [Internet]. arXiv; 2022 [cited 2023 Jul 24]. Available from: http://arxiv.org/abs/2204.00401

20. Goodfellow IJ, Pouget-Abadie J, Mirza M, Xu B, Warde-Farley D, Ozair S, et al. Generative Adversarial Networks. arXiv:14062661 [cs, stat] [Internet]. 2014 Jun 10 [cited 2021 May 27]; Available from: http://arxiv.org/abs/1406.2661

21. Papamakarios G, Nalisnick E, Rezende DJ, Mohamed S, Lakshminarayanan B. Normalizing Flows for Probabilistic Modeling and Inference [Internet]. arXiv; 2021 [cited 2023 Jul 24]. Available from: http://arxiv.org/abs/1912.02762

22. Qian Z, Cebere BC, van der Schaar M. Synthcity: facilitating innovative use cases of synthetic data in different data modalities [Internet]. arXiv; 2023 [cited 2023 Jul 24]. Available from: http://arxiv.org/abs/2301.07573

23. Chundawat VS, Tarun AK, Mandal M, Lahoti M, Narang P. TabSynDex: A Universal Metric for Robust Evaluation of Synthetic Tabular Data [Internet]. arXiv; 2022 [cited 2023 Jul 24]. Available from: http://arxiv.org/abs/2207.05295

24. Norcliffe A, Cebere B, Imrie F, Lio P, van der Schaar M. SurvivalGAN: Generating Time-to-Event Data for Survival Analysis [Internet]. arXiv; 2023 [cited 2023 Aug 3]. Available from: http://arxiv.org/abs/2302.12749

25. Platzer M, Reutterer T. Holdout-Based Fidelity and Privacy Assessment of Mixed-Type Synthetic Data [Internet]. arXiv; 2021 [cited 2023 Aug 10]. Available from: http://arxiv.org/abs/2104.00635

26. Hamming RW. Error detecting and error correcting codes. The Bell System Technical Journal. 1950 Apr;29(2):147–60.

27. Shuster JJ. Median follow-up in clinical trials. J Clin Oncol. 1991 Jan;9(1):191–2.

28. Emam KE, Jonker E, Arbuckle L, Malin B. A Systematic Review of Re-Identification Attacks on Health Data. PLOS ONE. 2011 Dec 2;6(12):e28071.

29. Ursin G, Sen S, Mottu JM, Nygård M. Protecting Privacy in Large Datasets-First We Assess the Risk; Then We Fuzzy the Data. Cancer Epidemiol Biomarkers Prev. 2017 Aug 1;26(8):1219–24.

30. Sweeney L, Yoo JS, Perovich L, Boronow KE, Brown P, Brody JG. Re-identification Risks in HIPAA Safe Harbor Data: A study of data from one environmental health study. Technol Sci. 2017;2017:2017082801.

31. Rieke N, Hancox J, Li W, Milletarì F, Roth HR, Albarqouni S, et al. The future of digital health with federated learning. npj Digit Med. 2020 Sep 14;3(1):1–7.

32. Warnat-Herresthal S, Schultze H, Shastry KL, Manamohan S, Mukherjee S, Garg V, et al. Swarm Learning for decentralized and confidential clinical machine learning. Nature. 2021 Jun;594(7862):265–70.

33. Melis L, Song C, De Cristofaro E, Shmatikov V. Exploiting Unintended Feature Leakage in Collaborative Learning [Internet]. arXiv; 2018 [cited 2023 Jul 10]. Available from: http://arxiv.org/abs/1805.04049

34. Zhu L, Liu Z, Han S. Deep Leakage from Gradients [Internet]. arXiv; 2019 [cited 2023 Jul 10]. Available from: http://arxiv.org/abs/1906.08935

35. Boenisch F, Dziedzic A, Schuster R, Shamsabadi AS, Shumailov I, Papernot N. When the Curious Abandon Honesty: Federated Learning Is Not Private [Internet]. arXiv; 2023 [cited 2023 Jul 10]. Available from: http://arxiv.org/abs/2112.02918

36. Rajotte JF, Bergen R, Buckeridge DL, El Emam K, Ng R, Strome E. Synthetic data as an enabler for machine learning applications in medicine. iScience. 2022 Nov 18;25(11):105331.

37. Chen RJ, Lu MY, Chen TY, Williamson DFK, Mahmood F. Synthetic data in machine learning for medicine and healthcare. Nat Biomed Eng. 2021 Jun;5(6):493–7.

38. Azizi Z, Zheng C, Mosquera L, Pilote L, Emam KE. Can synthetic data be a proxy for real clinical trial data? A validation study. BMJ Open. 2021 Apr 1;11(4):e043497.

39. D’Amico S, Dall’Olio D, Sala C, Dall’Olio L, Sauta E, Zampini M, et al. Synthetic Data Generation by Artificial Intelligence to Accelerate Research and Precision Medicine in Hematology. JCO Clinical Cancer Informatics. 2023 Jul;(7):e2300021.

