## Supplements for "Mimicking Clinical Trials with Synthetic Acute Myeloid Leukemia Patients Using Generative Artificial Intelligence"

| patient variable | data type |
|---|---|
| **demographic/clinical** | |
| age | continuous |
| sex | binary |
| AML status (de novo, sAML, tAML) | categorical |
| extramedullary manifestations | binary |
| **laboratory values** | |
| white blood cell count | continuous |
| hemoglobin level | continuous |
| platelet count | continuous |
| **outcome** | |
| achievement of CR | binary |
| EFS duration | continuous |
| EFS status | binary |
| OS duration | continuous |
| OS status | binary |
| **molecular genetics** | |
| ASXL1 | binary |
| BCOR | binary |
| BCORL1 | binary |
| DNTM3A | binary |
| EZH2 | binary |
| IDH1 | binary |
| IDH2 | binary |
| TET2 | binary |
| RAD21 | binary |
| SMC1A | binary |
| SMC3 | binary |
| STAG2 | binary |
| CEBPA | binary |
| CEBPA-bZIP in frame | binary |
| CUX1 | binary |
| GATA2 | binary |
| IKZF1 | binary |
| PHF6 | binary |
| RUNX1 | binary |
| WT1 | binary |
| TP53 | binary |
| NPM1 | binary |
| CBL | binary |
| CSF3R | binary |
| FLT3-ITD | binary |
| FLT3-TKD | binary |
| JAK2 | binary |
| KIT | binary |
| KRAS | binary |
| NOTCH1 | binary |
| NRAS | binary |
| PTPN11 | binary |
| SF3B1 | binary |

| | |
|---|---|
| SRSF2 | binary |
| U2AF1 | binary |
| ZRSR2 | binary |
| **cytogenetics** | |
| normal karyotype | binary |
| complex karyotype | binary |
| t(8;21) | binary |
| inv(16) or t(16;16) | binary |
| t(6;9) | binary |
| inv(3) or t(3;3) | binary |
| t(9;11) | binary |
| t(v;11) | binary |
| t(9;22) | binary |
| -5 | binary |
| del(5q) | binary |
| -7 | binary |
| del(7q) | binary |
| -17 or del(17p) | binary |
| abn(17p) | binary |

**Table S1 Available patient variables included in synthetic data generation.** Abbreviations: complete remission (CR), event-free survival (EFS), overall survival (OS), secondary acute myeloid leukemia (sAML), therapy-associated acute myeloid leukemia (tAML).

| trial name | clinicaltrials.gov identifier | trial duration | protocol summary |
|---|---|---|---|
| AML96 | NCT00180115 | 1996-2008 | risk-adapted postremission treatment regarding allogeneic stem cell transplantation for high-risk AML and related allogeneic and autologous stem cell transplantation for standard-risk AML, and randomization between intermediate-dose and high-dose cytarabine within the first post-remission course |
| AML2003 | NCT00180102 | 2003-2009 | early allogeneic stem cell transplantation in post-induction aplasia for high-risk AML, factorial design with four therapy arms with two factors of two stages (intensified vs. standard therapy and |

| | | | |
|---|---|---|---|
| | | | cytarabine vs. cytarabine + mitoxantrone + amsacrin) |
| AML60+ | NCT00180167 | 2005-2010 | Patients ≥ 60 years, mitoxantron on day 1,2,3 + cytarabine on days 1,3,5,7 vs. DA 7+3 |
| SORAML | NCT00893373 | 2011-2014 | Standard therapy + sorafenib vs. standard therapy + placebo |

**Table S1.** Summary of trial regimens

| **Performance metric** | Explanation | Reference |
|---|---|---|
| **Basic Statistical Measure** | compares the mean, median, and standard deviation between numerical columns of both real and synthetic datasets to assess their similarity | Chundawat et al. |
| **Regularized Support Coverage** | quantifies the overlap in individual feature distributions between the original and synthetic datasets, ensuring both share similar support for each feature | Chundawat et al. |
| **Log-transformed Correlation Score** | evaluates the difference in correlation matrices between the original and synthetic datasets, which helps assess how well the synthetic data captures inter-feature relationships | Chundawat et al. |

| | | |
|---|---|---|
| **Kaplan-Meier Divergence** | This metric calculates the mean absolute difference between the Kaplan-Meier survival curves of the synthetic and real data, measuring the overall match between the survival probabilities. | Norcliffe et al. |
| **Optimism** | This survival analysis metric measures the discrepancy in expected lifetimes between the synthetic and real data, as illustrated by their respective Kaplan-Meier survival curves. It quantifies the degree of over-optimism or over-pessimism in the synthetic data | Norcliffe et al. |
| **Short-Sightedness** | This metric quantifies the extent to which models, as evaluated by Kaplan-Meier survival curves of synthetic data, fail to predict beyond a certain time horizon, capturing temporal limitations in the synthetic data | Norcliffe et al. |

**Table S3 Performance metrics for fidelity and usability of synthetic data**.

| performance metric | original cohort | CTAB-GAN+ | NFLOW |
|---|---|---|---|
| Log-transformed Correlation Score | 0.58 | 0.75 | 0.74 |
| Regularized Support Coverage | 0.93 | 0.95 | 0.97 |
| Basic Statistical Measure | 0.95 | 0.91 | 0.92 |
| Optimism | 1.00 | 0.97 | 0.98 |
| Kaplan-Meier Divergence | 1.00 | 0.98 | 0.99 |
| Short-Sightedness | 0.94 | 0.99 | 0.93 |

**Table S4 Performance evaluation of both generative models.** Previously proposed performance metrics for tabular synthetic data (introduced by Chundawat et al.[22] and Norcliffe et al.[23]) were used to evaluate model performance. All metrics are scaled from 0 (inadequate representation of original data) to 1 (optimal representation).

| | | original cohort | CTAB-GAN+ | *p* | NFlow | *p* |
|---|---|---|---|---|---|---|
| number of patients | | 1606 | 1606 | | 1606 | |
| **molecular genetics, n(%)** | | | | | | |
| epigenetic | *ASXL1* | 126 (7.9) | 161 (10.0) | 0.035 | 113 (7.0) | 0.420 |
| | *BCOR* | 76 (4.7) | 78 (4.9) | 0.934 | 72 (4.5) | 0.801 |
| | *BCORL1* | 60 (3.7) | 59 (3.7) | 1.000 | 182 (11.3) | 0.000 |
| | *DNTM3A* | 458 (28.5) | 413 (25.7) | 0.081 | 547 (34.0) | 0.001 |
| | *EZH2* | 63 (3.9) | 75 (4.7) | 0.339 | 63 (3.9) | 1.000 |
| | *IDH1* | 149 (9.3) | 156 (9.7) | 0.718 | 149 (9.3) | 1.000 |
| | *IDH2* | 227 (14.1) | 214 (13.3) | 0.538 | 278 (17.3) | 0.015 |

| | | | | | | |
|---|---|---|---|---|---|---|
| | *TET2* | 311 (19.4) | 313 (19.5) | 0.964 | 308 (19.2) | 0.929 |
| transcription | *CEBPA* | 257 (16.0) | 323 (20.1) | 0.002 | 268 (16.7) | 0.503 |
| | *CEBPA*. biallelic | 92 (5.7) | 132 (8.2) | 0.001 | 89 (5.5) | 0.878 |
| | *CEBPA*-TAD | 37 (2.3) | 50 (3.1) | 0.102 | 24 (1.5) | 0.194 |
| | *CEBPA*-bZIP | 144 (9.0) | 206 (12.8) | 0.851 | 127 (7.9) | 0.001 |
| | *CUX1* | 44 (2.7) | 86 (5.4) | <0.001 | 48 (3.0) | 0.751 |
| | *GATA2* | 97 (6.0) | 159 (9.9) | <0.001 | 109 (6.8) | 0.428 |
| | *IKZF1* | 45 (2.8) | 39 (2.4) | 0.581 | 37 (2.3) | 0.434 |
| | *PHF6* | 52 (3.2) | 65 (4.1) | 0.258 | 109 (6.8) | 0.000 |
| | *RUNX1* | 147 (9.2) | 159 (9.9) | 0.509 | 156 (9.7) | 0.629 |
| | *WT1* | 118 (7.4) | 80 (5.0) | 0.007 | 117 (7.3) | 1.000 |
| signaling | *CBL* | 32 (2.0) | 44 (2.7) | 0.201 | 28 (1.7) | 0.696 |
| | *CSF3R* | 29 (1.8) | 44 (2.7) | 0.097 | 33 (2.1) | 0.701 |
| | *FLT3*-ITD | 349 (21.7) | 347 (21.6) | 1.000 | 363 (22.6) | 0.496 |
| | *FLT3*-TKD | 62 (3.9) | 53 (3.3) | 0.633 | 94 (5.9) | 0.004 |
| | *JAK2* | 18 (1.1) | 23 (1.4) | 0.530 | 22 (1.4) | 0.634 |
| | *KIT* | 79 (4.9) | 97 (6.0) | 0.187 | 75 (4.7) | 0.804 |
| | *KRAS* | 85 (5.3) | 115 (7.2) | 0.034 | 77 (4.8) | 0.573 |
| | *NOTCH1* | 32 (2.0) | 39 (2.4) | 0.472 | 43 (2.7) | 0.242 |

| | | | | | | |
|---|---|---|---|---|---|---|
| | *NRAS* | 249 (15.5) | 305 (19.0) | 0.010 | 198 (12.3) | 0.011 |
| | *PTPN11* | 113 (7.0) | 119 (7.4) | 0.733 | 102 (6.4) | 0.480 |
| splicing | *SF3B1* | 46 (2.9) | 48 (3.0) | 0.917 | 41 (2.7) | 0.664 |
| | *SRSF2* | 102 (6.4) | 101 (6.3) | 1.000 | 138 (8.6) | 0.019 |
| | *U2AF1* | 45 (2.8) | 51 (3.2) | 0.605 | 45 (2.8) | 1.000 |
| | *ZRSR2* | 26 (1.6) | 20 (1.3) | 0.458 | 64 (4.0) | 0.000 |
| cohesin | *RAD21* | 51 (3.2) | 58 (3.6) | 0.559 | 44 (2.7) | 0.532 |
| | *SMC1A* | 23 (1.4) | 26 (1.6) | 0.774 | 22 (1.4) | 1.000 |
| | *SMC3* | 18 (1.1) | 31 (1.9) | 0.083 | 37 (2.3) | 0.014 |
| | *STAG2* | 88 (5.5) | 69 (4.3) | 0.141 | 132 (8.2) | 0.003 |
| other | *TP53* | 114 (7.1) | 100 (6.2) | 0.358 | 115 (7.2) | 1.000 |
| | *NPM1* | 501 (31.2) | 507 (31.6) | 0.819 | 508 (31.6) | 0.674 |
| **cytogenetics. n (%)** | | | | | | |
| | normal karyotype | 830 (51.7) | 780 (48.6) | 0.085 | 788 (49.1) | 0.143 |
| | complex karyotype | 188 (11.7) | 209 (13.0) | 0.280 | 229 (14.3) | 0.072 |

| | | | | | |
|---|---|---|---|---|---|
| t(8;21) | 61 (3.8) | 90 (5.6) | 0.019 | 71 (4.4) | 0.424 |
| inv(16) or t(16;16) | 101 (6.3) | 101 (6.3) | 1.000 | 89 (5.5) | 0.371 |
| t(6;9) | 6 (0.4) | 5 (0.3) | 0.774 | 11 (0.7) | 0.331 |
| inv(3) or t(3;3) | 7 (0.4) | 17 (1.1) | 0.063 | 10 (0.6) | 0.628 |
| t(9;11) | 11 (0.7) | 22 (1.4) | 0.079 | 22 (1.4) | 0.079 |
| t(v;11) | 16 (1.0) | 34 (2.1) | 0.015 | 15 (0.9) | 0.859 |
| t(9;22) | 3 (0.2) | 3 (0.2) | 1.000 | 12 (0.8) | 0.035 |
| -5 | 24 (1.5) | 28 (1.7) | 0.675 | 31 (1.9) | 0.415 |
| del(5q) | 18 (1.1) | 23 (1.4) | 0.530 | 20 (1.3) | 0.871 |
| -7 | 71 (4.4) | 83 (5.2) | 0.364 | 92 (5.7) | 0.108 |
| del(7q) | 16 (1.0) | 15 (0.9) | 0.859 | 36 (2.2) | 0.007 |
| -17 | 34 (2.1) | 38 (2.4) | 0.721 | 21 (1.3) | 0.079 |
| abn(17p) | 6 (0.4) | 13 (0.8) | 0.166 | 18 (1.1) | 0.022 |

**Table S5 Distribution of molecular and cytogenetic alterations between the original and the synthetic cohorts.** *p*-values are calculated using two-sample comparisons between each of the synthetic cohorts and the baseline cohort for reference. Abbreviations: number (n).

| | **original** | ***p*** | **CTAB-GAN+** | ***p*** | **NFlow** | ***p*** |
|---|---|---|---|---|---|---|
| age | 0.94 [0.93-0.95] | <0.001 | 0.94 [0.93-0.95] | <0.001 | 0.95 [0.94-0.95] | <0.001 |

| | | | | | | |
|---|---|---|---|---|---|---|
| normal karyotype | 1.98 [1.58-2.49] | <0.001 | 2.22 [1.75-2.81] | <0.001 | 1.50 [1.20-1.88] | <0.001 |
| complex karyotype | 0.40 [0.29-0.54] | <0.001 | 0.39 [0.29-0.53] | <0.001 | 0.58 [0.43-0.77] | <0.001 |
| inv(16) or t(16;16) | 3.25 [1.76-5.99] | <0.001 | 1.82 [1.07-3.10] | 0.028 | 2.73 [1.50-4.97] | 0.001 |
| t(8;21) | 8.38 [2.61-26.89] | <0.001 | 3.37 [1.68-6.77] | 0.001 | 3.20 [1.58-6.49] | 0.001 |
| t(9;11) | 1.87 [0.40-8.69] | 0.424 | 1.21 [0.45-3.31] | 0.704 | 0.78 [0.32-1.87] | 0.576 |
| -5 | 0.13 [0.05-0.34] | <0.001 | 0.16 [0.07-0.36] | <0.001 | 0.24 [0.11-0.50] | <0.001 |
| del(5q) | 0.33 [0.13-0.83] | 0.019 | 0.07 [0.02-0.21] | <0.001 | 0.67 [0.27-1.64] | 0.378 |
| -7 | 0.25 [0.15-0.41] | <0.001 | 0.24 [0.15-0.37] | <0.001 | 0.20 [0.13-0.32] | <0.001 |
| -17 | 0.12 [0.05-0.27] | <0.001 | 0.10 [0.05-0.22] | <0.001 | 0.33 [0.14-0.79] | 0.013 |
| *NPM1* | 2.49 [1.91-3.24] | <0.001 | 2.80 [2.11-3.70] | <0.001 | 1.69 [1.33-2.15] | <0.001 |
| *FLT3*-ITD | 1.79 [1.35-2.39] | <0.001 | 2.12 [1.55-2.91] | <0.001 | 1.41 [1.08-1.84] | 0.011 |
| *CEBPA*-bZIP (inframe) | 8.12 [3.56-18.57] | <0.001 | 4.88 [2.47-9.67] | <0.001 | 3.56 [1.78-7.15] | <0.001 |
| *TP53* | 0.14 [0.09-0.22] | <0.001 | 0.17 [0.11-0.26] | <0.001 | 0.17 [0.11-0.26] | <0.001 |

| | | | | | | |
|---|---|---|---|---|---|---|
| *RUNX1* | 0.30 [0.21-0.42] | <0.001 | 0.20 [0.14-0.28] | <0.001 | 0.54 [0.39-0.76] | <0.001 |
| *ASXL1* | 0.35 [0.25-0.51] | <0.001 | 0.42 [0.30-0.58] | <0.001 | 0.46 [0.31-0.68] | <0.001 |

**Table S6 Comparative univariable analyses for individual patient variables with respect to achievement of complete remission.** Variables with previously demonstrated impact on patient outcome were analyzed using univariable logistic regression. Their odds ratio (OR) and 95%-confidence interval (square brackets) as well as corresponding *p*-values are reported per cohort. Except for del(5q) being significantly associated with failure to achieve CR in the original cohort while this effect turned out to be non-significant in the NFlow-generated cohort, all other effects were of the same directionality and statistical significance. Importantly, no variable showed an inverted effect (for example, a favorable marker turning unfavorable in a synthetic cohort).

| | **original** | ***p*** | **CTAB-GAN+** | ***p*** | **NFlow** | ***p*** |
|---|---|---|---|---|---|---|
| age | 1.03 [1.03-1.03] | <0.001 | 1.03 [1.03-1.03] | <0.001 | 1.03 [1.03-1.04] | <0.001 |
| normal karyotype | 0.82 [0.73-0.93] | 0.001 | 0.85 [0.76-0.96] | 0.008 | 0.82 [0.73-0.92] | 0.001 |
| complex karyotype | 1.64 [1.39-1.93] | <0.001 | 1.68 [1.43-1.98] | <0.001 | 1.44 [1.23-1.69] | <0.001 |
| inv(16) or t(16;16) | 0.58 [0.44-0.74] | <0.001 | 0.43 [0.32-0.58] | <0.001 | 0.58 [0.44-0.77] | <0.001 |
| t(8;21) | 0.35 [0.21-0.52] | <0.001 | 0.38 [0.28-0.53] | <0.001 | 0.41 [0.29-0.58] | <0.001 |
| t(9;11) | 0.64 [0.30-1.34] | 0.237 | 0.84 [0.50-1.42] | 0.525 | 1.19 [0.74-1.92] | 0.481 |

| | | | | | | |
|---|---|---|---|---|---|---|
| -5 | 3.53 [2.35-5.30] | <0.001 | 3.92 [2.68-5.72] | <0.001 | 2.51 [1.72-3.69] | <0.001 |
| del(5q) | 2.76 [1.73-4.41] | <0.001 | 3.40 [2.24-5.15] | <0.001 | 1.76 [1.10-2.80] | 0.018 |
| -7 | 2.82 [2.20-3.61] | <0.001 | 3.10 [2.47-3.90] | <0.001 | 2.97 [2.37-3.71] | <0.001 |
| -17 | 3.32 [2.34-4.70] | <0.001 | 3.50 [2.42-4.84] | <0.001 | 1.97 [1.23-3.13] | 0.004 |
| *NPM1* | 0.68 [0.60-0.77] | <0.001 | 0.77 [0.68-0.87] | <0.001 | 0.73 [0.64-0.82] | <0.001 |
| *FLT3*-ITD | 1.00 [0.88-1.15] | 0.959 | 1.04 [0.91-1.20] | 0.564 | 0.95 [0.83-1.09] | 0.461 |
| *CEBPA*-bZIP (inframe) | 0.39 [0.28-0.54] | <0.001 | 0.44 [0.34-0.60] | <0.001 | 0.64 [0.47-0.88] | 0.006 |
| *TP53* | 2.82 [2.31-3.44] | <0.001 | 3.34 [2.71-4.13] | <0.001 | 3.05 [2.50-3.73] | <0.001 |
| *RUNX1* | 1.88 [1.57-2.24] | <0.001 | 1.95 [1.63-2.31] | <0.001 | 1.76 [1.48-2.10] | <0.001 |
| *ASXL1* | 1.86 [1.54-2.25] | <0.001 | 1.62 [1.31-2.01] | <0.001 | 1.52 [1.28-1.81] | <0.001 |

**Table S7 Comparative univariable analyses for individual patient variables with respect to event-free survival.** Variables with previously demonstrated impact on patient outcome were analyzed using univariable logistic regression. Their Hazard Ratio (HR) and 95%-confidence interval (square brackets) as well as corresponding *p*-values are reported per cohort. No discrepancies between effect direction and statistical significances of effects were found between the original and both synthetic cohorts were found.

| | original | *p* | CTAB-GAN+ | *p* | NFlow | *p* |
|---|---|---|---|---|---|---|
| age | 1.04<br>[1.03-1.04] | <0.001 | 1.03<br>[1.03-1.04] | <0.001 | 1.04<br>[1.03-1.05] | <0.001 |
| normal karyotype | 0.80<br>[0.71-0.91] | 0.001 | 0.78<br>[0.69-0.88] | <0.001 | 0.81<br>[0.71-0.92] | 0.001 |
| complex karyotype | 1.72<br>[1.44-2.04] | <0.001 | 1.86<br>[1.58-2.20] | <0.001 | 1.42<br>[1.20-1.67] | <0.001 |
| inv(16) or t(16;16) | 0.52<br>[0.39-0.70] | <0.001 | 0.41<br>[0.29-0.57] | <0.001 | 0.56<br>[0.41-0.78] | <0.001 |
| t(8;21) | 0.33<br>[0.21-0.51] | <0.001 | 0.37<br>[0.26-0.53] | <0.001 | 0.41<br>[0.28-0.60] | <0.001 |
| t(9;11) | 0.60<br>[0.25-1.43] | 0.247 | 0.88<br>[0.50-1.55] | 0.655 | 1.22<br>[0.74-2.04] | 0.436 |
| -5 | 4.37<br>[2.90-6.57] | <0.001 | 4.30<br>[2.95-6.28] | <0.001 | 2.44<br>[1.66-3.57] | <0.001 |
| del(5q) | 2.32<br>[1.41-3.80] | 0.001 | 3.22<br>[2.13-4.88] | <0.001 | 1.83<br>[1.14-2.96] | 0.013 |
| -7 | 2.79<br>[2.17-3.58] | <0.001 | 3.16<br>[2.41-3.97] | <0.001 | 2.77<br>[2.21-3.46] | <0.001 |

| | | | | | | |
|---|---|---|---|---|---|---|
| -17 | 3.68 [2.59-5.21] | <0.001 | 3.46 [2.49-4.79] | <0.001 | 1.91 [1.19-3.09] | 0.008 |
| *NPM1* | 0.74 [0.65-0.85] | <0.001 | 0.72 [0.65-0.82] | <0.001 | 0.75 [0.65-0.86] | <0.001 |
| *FLT3*-ITD | 1.06 [0.92-1.22] | 0.440 | 1.01 [0.87-1.18] | 0.853 | 0.95 [0.82-1.10] | 0.468 |
| *CEBPA*-bZIP (inframe) | 0.41 [0.28-0.59] | <0.001 | 0.42 [0.31-0.57] | <0.001 | 0.68 [0.48-0.96] | 0.027 |
| *TP53* | 3.44 [2.81-4.21] | <0.001 | 3.65 [2.85-4.52] | <0.001 | 2.75 [2.24-3.37] | <0.001 |
| *RUNX1* | 1.82 [1.51-2.19] | <0.001 | 1.92 [1.61-2.30] | <0.001 | 1.69 [1.41-2.03] | <0.001 |
| *ASXL1* | 1.64 [1.35-2.01] | <0.001 | 1.55 [1.29-1.87] | <0.001 | 1.68 [1.36-2.08] | <0.001 |

**Table S8 Comparative univariable analyses for individual patient variables with respect to overall survival.** Variables with previously demonstrated impact on patient outcome were analyzed using univariable logistic regression. Their Hazard Ratio (HR) and 95%-confidence interval (square brackets) as well as corresponding *p*-values are reported per cohort. No discrepancies between effect direction and statistical significances of effects were found between the original and both synthetic cohorts were found.

| | CTAB-GAN+ | NFlow | original cohort |
|---|---|---|---|
| **absolute Hamming distances** | | | |
| average min. distance train | 8.7034 | 9.3474 | 8.2524 |
| average min. distance test | 8.8587 | 9.4117 | 8.2224 |
| median distance train | 9 | 9 | 8 |
| median distance test | 9 | 9 | 8 |
| **relative Hamming distances** | | | |
| privacy leakage coefficient | 0.0178 | 0.0069 | |

**Table S9 Hamming distances for privacy conservation**. Hamming distances were used to measure the distance between two points within and between equally sized subsets of training (four sets of 20%) and test data (20%). The median distance represents the number of variables that have to be altered (and matched exactly) to fit a real patient. A threshold for the privacy leakage coefficient of 0.05 for relative distances was set where values above 0.05 signal potential privacy breaches. Both synthetic data sets fell well below the 0.05 threshold signaling larger distances between synthetic and training data.

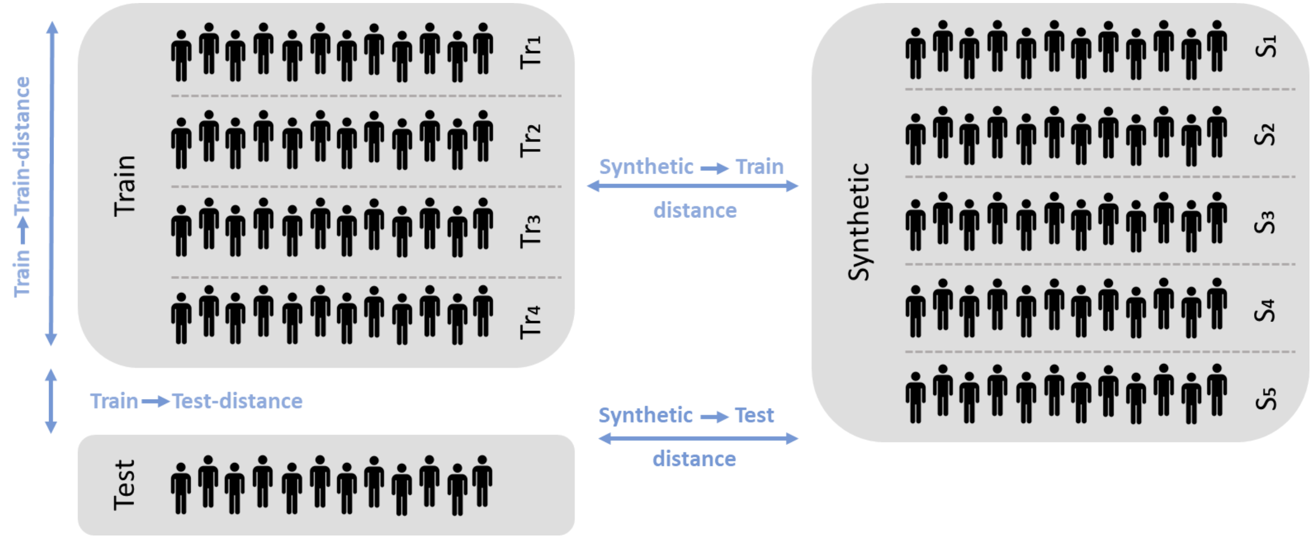

**Figure S1 Partitioning of privacy assessment subsets.** Because of the mismatch between training set size (80% of the total cohort) and test set size (20%), both the training set and the synthetic cohort were partitioned into equally sized (20% each) subsets in order to guarantee adequate comparability via Hamming distance calculation.
